# Prediction of 5-year mortality risk in 784,892 people with mental illness: Development and validation of a novel clinical prognostic model (MortOx)

**DOI:** 10.64898/2025.12.17.25342475

**Authors:** Amir Sariaslan, Thomas Fanshawe, Jonas Forsman, Joonas Pitkänen, Ralf Kuja-Halkola, Isabell Brikell, Zheng Chang, Henrik Larsson, Pekka Martikainen, Paul Lichtenstein, Seena Fazel

## Abstract

Individuals with psychiatric disorders have significantly higher mortality rates than the general population. Despite identifying risk factors, few attempts have been made to systematically use this information to stratify mortality risk. To address this gap, we developed and externally validated a risk prediction model using national healthcare and social register data from two countries. Nationwide register data were used to create a Swedish cohort (n=530,201) for model development and a Finnish cohort (n=254,691) for external validation. Participants, aged 15-60 years at assessment, had diagnoses of common mental illnesses (schizophrenia-spectrum disorder, bipolar disorder, depression, or anxiety disorders) made in specialist care. A multivariable logistic regression model assessed predictors of 5-year mortality risk. Model performance was evaluated using discrimination (area under the curve [AUC]) and calibration metrics, including intercept, slope, and visual plots. Internal validation employed bootstrapping. A total of 18,619 (3.5%) Swedish and 11,206 (4.4%) Finnish patients died within 5 years of assessment in secondary care. The model incorporated 25 predictors across four domains: sociodemographic factors, clinical characteristics, somatic comorbidities, and psychotropic medication use. External validation demonstrated excellent discrimination (AUC = 0.803; 95% CI: 0.799-0.807). The higher mortality rate in the Finnish cohort required recalibration of the intercept, and post-adjustment calibration was good (intercept: 0.00; 95% CI: −0.02; 0.02; slope: 0.99; 95% CI: 0.98-1.01). Model findings were translated into a web-based calculator (MortOx) for research, training, and potential clinical use. A transdiagnostic prognostic model based on 25 predictors accurately predicts 5-year mortality risk in mental illness. Linkage to interventions is needed to evaluate clinical impact.

---

Preventing premature mortality in people with mental illness remains a pressing public health priority,^1^ with estimates of the potential life years lost in this population ranging between 7 and 22 years across specific disorders.^2,3^ Epidemiological studies have identified several risk factors contributing to this excess mortality, including co-occurring substance use disorders,^4^ comorbid somatic conditions,^4,5^ psychotropic medication nonadherence,^6^ and familial risk of premature mortality.^5^ However, it remains unclear to what extent these risk factors can be used to develop prognostic models and tools to accurately stratify mortality risk in this population. Such models have been developed for the general population^7^ and other areas of medicine, the latter of which being widely used to support clinical decision-making and optimise resource allocation.^8,9^

A recent systematic review^10^ identified several tools developed to predict suicidal outcomes, but none that specifically focus on all-cause mortality. This is an important gap in the literature, as previous reports suggest that suicide and other unnatural causes of death account for only up to a quarter of all deaths among people with mental illness.^11,12^ Importantly, the increasing mortality gap between people with severe mental illness and the general population is largely driven by excess life years lost due to cardiovascular and respiratory diseases.^13–15^ Although many risk factors for suicide and all-cause mortality overlap, there are important differences in their relative effect sizes,^2^ which suggests that tools to stratify suicide risk may not perform as well for all-cause mortality.

To address this evidence gap, we aimed to develop and validate a scalable, transdiagnostic prognostic tool for predicting 5-year mortality risk in patients with common mental illnesses (e.g., psychotic disorders, depression, and anxiety). We used routinely collected data from Swedish and Finnish population healthcare and other registers to allow for sufficient statistical power, sample representativeness, and reliability for predictors and outcome, and also external validation in a new country. We evaluated the performance of the risk model across specific geographic regions and sociodemographic groups, as mortality gradients by sociodemographic groups have been well established.^13,16^ The model is intended for use by researchers and clinicians.

## Methods

All Swedish and Finnish residents receive a unique personal identification number at birth or upon immigration, allowing accurate linkage across multiple national social and health registers.^17^ We obtained access to pseudonymised administrative population data from the Swedish Ethical Review Authority (Dnr 2020-06540; Dnr 2022-06204-02), the Ethics Board of Statistics Finland (TK-53-1490-18), and the Finnish Institute of Health and Welfare (THL/2180/14.02.00/2020). Under Swedish and Finnish legislation, informed consent is not required for research using national registers with pseudonymised identifiers. We followed the TRIPOD+AI guidelines.^18^

We generated the development/derivation sample by combining data from the Swedish population registers and the National Patient Register.^19^ This allowed us to identify individuals born in Sweden, aged 15 to 60 years, who could be linked to their biological parents, and who had received at least one secondary care diagnosis (i.e. excluding solely primary care diagnoses) of a schizophrenia-spectrum disorder, bipolar disorder, depression, or anxiety (using a hierarchical classification; ICD codes are listed in **eTable 1**) between 2006 and 2016 (see **eFigure 1** for the sample flowchart). If individuals had multiple mental illnesses or episodes during this period, we randomly selected one episode to ensure that the model would be applicable to any patient episode. For external validation, we included a similar cohort in Finland from The Care Register for Health Care^20^ between 2006 and 2014. Both registers contained nearly all inpatient hospitalisations and outpatient specialist visits during these time frames, although data on treatment delivered during each episode were unavailable, with the exception of recently prescribed medications. Statistical power was excellent across both samples (**eText 1**).

### Outcome

We obtained data on mortality dates, including both primary and contributory causes of death, from the Causes of Death Registers in each country.^21^ In Sweden, coroners’ reports had a coverage rate exceeding 99% from 2012 to 2020.^22^ In Finland, it is legally mandated for the cause of cause of death to be ascertained before an individual can be buried (Act on Determining the Cause of Death 459/1973). The primary outcome was defined as death from any cause within five years of assessment. For descriptive purposes, we further categorised the causes of death as suicides, other external causes, or natural causes (ICD codes listed in **eTable 1**).

### Candidate predictors

A total of 31 candidate predictors were selected, based on a review of the literature,^2,4,5,23^ and harmonised across both cohorts. These covered five domains: sociodemographic factors, familial risk, clinical characteristics, somatic comorbidities, and recent psychotropic medication treatments prescribed prior to the assessment (detailed in **Supplementary Materials**). Interaction terms between recent psychotropic medications and mental illnesses were pre-specified, based on the assumption that medication effects might vary by diagnosis.

### Model development

We fitted a multivariable logistic regression model to estimate the associations between candidate predictors, including all pre-specified interaction terms, and all-cause mortality within five years of assessment. Predictor selection was conducted using backward stepwise elimination on the basis of Akaike’s Information Criterion.^24^ Age at assessment was modelled as a continuous variable (in units of decades), with linearity supported by visual inspection of its smoothed partial residual plot. Although we initially considered multiple imputation to address missing data, the low percentage of missingness (<5%), in addition to the computational demands of downstream analyses required for internal validation, led us to adopt listwise deletion (e.g., complete-case analysis) for the main results. However, estimates based on multiple imputation are provided for comparison.

### Internal validation

Internal validation involved assessing performance metrics to evaluate the discriminatory accuracy and calibration of the model using the Swedish development dataset. Discriminatory accuracy measures the ability of the model to distinguish between individuals who died within five years and those who survived. Calibration, on the other hand, tests the agreement between predicted and observed mortality risks.

Discriminatory accuracy was quantified using the area under the receiver operating characteristics (ROC) curve (AUC), which ranges from 0.5 (no better than chance) to 1.0 (perfect discrimination). Calibration was evaluated using two primary metrics: the calibration intercept (calibration-in-the-large; CITL) and the calibration slope. To visualise the agreement between predicted and observed risks, calibration plots were generated based on decile groupings and smoothed calibration curves using generalised additive modelling techniques. These plots provided a visual representation of calibration across the entire range of predicted risks. Additional metrics, such as the Integrated Calibration Index (ICI), were calculated to provide a summary measure of miscalibration. The ICI represents the weighted average of absolute differences between the smoothed calibration curve and perfect calibration, with weights based on the predicted risk distribution. Furthermore, the 90th percentile (E90) and maximum (Emax) absolute differences were calculated to evaluate the extent of potential miscalibration across the full range of predicted risks.

To address optimism or overfitting, which occurs when a model performs well on the derivation dataset but suboptimally on external validation, we used a bootstrap approach.^25^ In brief, we resampled the derivation dataset 200 times with replacement and applied the same analytical procedures that were used to fit the final model, thus allowing for variations in predictor inclusion across resamples. Optimism was subsequently calculated as the difference between the average performance metrics (e.g., AUC, calibration intercept, and slope) across the bootstrap models and the corresponding estimates from the original derivation dataset. Correcting for optimism increases the likelihood that the predictions will generalise to independent datasets.

In addition to correcting for optimism, the bootstrap approach was also used to evaluate the stability of predicted risks across varying population compositions (based on resampling of the derivation sample) and predictor sets. Variation in individual-level predictions between the final model and bootstrapped models was summarised using a 95% stability interval, defined as the range encompassing the middle 95% of predictions from the bootstrapped models.^26^ A stability plot was used to graphically assess the relationship between predicted risks from the derivation model and predictions from the bootstrapped models. Tight clustering of points in the stability plot indicates consistent predictions across models, thus suggesting robustness to variations in population composition and predictor inclusion.

We also assessed heterogeneity in model performance across all 21 counties in Sweden using an internal-external cross-validation approach.^25^ Five-year mortality risks for individuals in each county were predicted using models trained on data from all other counties. County-specific performance metrics were then pooled using random-effects meta-analysis. Heterogeneity was quantified using the τ^2^ statistic, representing between-county variance in absolute terms (in the same units as the underlying scale).

### External validation

To evaluate the performance of the model in the external validation sample of Finnish individuals, we applied the coefficients from the final derivation model to calculate individual-level predictions of five-year mortality risks. To account for differences in mortality rates between the countries, we conservatively recalibrated the intercept of the model using a recommended approach. ^27^ Furthermore, we conducted stratified analyses to evaluate differences in model performance across relevant subgroups defined by geographic region, sex, age categories (15-24, 25-29, 30-39, 40-49, and 50-60 years), and socioeconomic status (benefit receipt). These subgroup analyses aimed to assess fairness in model performance across diverse populations. We additionally tested for the model performance across clinical characteristics, including diagnostic category (e.g., anxiety, depression, bipolar disorder and schizophrenia-spectrum disorders), alternative time horizons (e.g., 1 and 2 years), and cause-specific mortality (e.g., natural and external causes, including substance and suicide related deaths).

The final model has been implemented in an online calculator, MortOx, that generates a probability score for 5-year all-cause mortality [URL to the tool be added].

### Patient and public involvement

We conducted a series of meetings with the Oxford Health BRC Data Science PPI group, which consists of individuals with lived experience of psychiatric disorders and, in some cases, comorbid physical illnesses. These meetings informed the refinement of our research questions, selection of predictors, and design of complementary subgroup analyses. Following the presentation of the final findings to the group, participants provided feedback on how they preferred the tool to be used in clinical settings and offered input on approaches to communicating the results (**eText 2**).

## Results

A total of 784,892 individuals (mean [SD] age at assessment: 37 [13] years) were included in the analysis, comprising 530,201 persons (37 [13] years) in the derivation sample and 254,691 (36 [14] years) in the external validation sample (eFigure 1). Over a five-year follow-up period, 18,619 (3.5%) individuals died in the derivation sample, compared with 11,206 (4.4%) deaths in the external validation sample. Mortality rates varied notably by diagnostic category, ranging from 2.2%-2.4% in anxiety to 7.8-9.2% in schizophrenia-spectrum disorders (**Table 1**). Fewer than a third (23.8%-28.6%) of deaths were attributed to suicide, with approximately an additional fifth (16.9% to 20.6%) resulting from other external causes (e.g., accidents and substance overdoses) across all included disorders (**eFigure 2**).

**Table 1.** Mortality rates (percentage) across the diagnostic categories in the derivation and external validation samples.

| | Number of individuals | Male sex, n (%) | Age at assessment, mean ± SD | Deaths within 5 years | |
| --- | --- | --- | --- | --- | --- |
| ***Derivation sample*** |  |  |  | **Events** | **Percent [95% CI]** |
| All individuals | 530,201 | 217,426 (41.0%) | 37 ± 13 | 18,619 | 3.5% [3.5%; 3.6%] |
| Anxiety | 213,878 | 85,297 (39.9%) | 35 ± 13 | 5112 | 2.4% [2.3%; 2.5%] |
| Depression | 233,368 | 92,494 (39.6%) | 36 ± 13 | 7788 | 3.3% [3.3%; 3.4%] |
| Bipolar disorder | 36,076 | 13,570 (37.6%) | 39 ± 13 | 1410 | 3.9% [3.7%; 4.1%] |
| Schizophrenia-spectrum disorder | 46,879 | 26,065 (55.6%) | 43 ± 12 | 4309 | 9.2% [8.9%; 9.5%] |
| ***External validation sample*** |  |  |  |  |  |
| All individuals | 254,691 | 105,873 (41.6%) | 36 ± 14 | 11,206 | 4.4% [4.3%; 4.5%] |
| Anxiety | 43,816 | 17,545 (40.0%) | 31 ± 13 | 953 | 2.2% [2.0%; 2.3%] |
| Depression | 140,767 | 52,418 (37.2%) | 35 ± 14 | 5111 | 3.6% [3.5%; 3.7%] |
| Bipolar disorder | 19,317 | 8417 (43.6%) | 39 ± 12 | 1183 | 6.1% [5.8%; 6.5%] |
| Schizophrenia-spectrum disorder | 50,791 | 27,493 (54.1%) | 40 ± 13 | 3959 | 7.8% [7.5%; 8.0%] |

The magnitude of the associations between the predictors and outcome was negligbly different when we either excluded missing data (i.e., complete-case analysis) or imputed them (**Table 2; eTable 2**). The final model retained 25 of the 31 predictors, including all interaction terms between diagnostic categories and recent psychotropic medications, except for those with antidepressants and psychostimulants. The distribution of the included predictors is shown in **Table 3**. Compared to the Swedish derivation sample, we found that individuals in the Finnish validation sample had lower levels of educational attainment (26.4% lacked secondary school qualifications in the derivation sample vs. 36.8% in the validation sample) and higher rates of benefit receipt (28.0% vs. 37.7%). Differences were also observed in the distribution of diagnostic categories, with the derivation sample including a larger proportion of individuals with anxiety disorders (40.3% vs. 17.2%) but smaller proportions of those with depression (44.0% vs. 55.3%) and schizophrenia-spectrum disorders (8.8% vs. 19.9%).

**Table 2.**
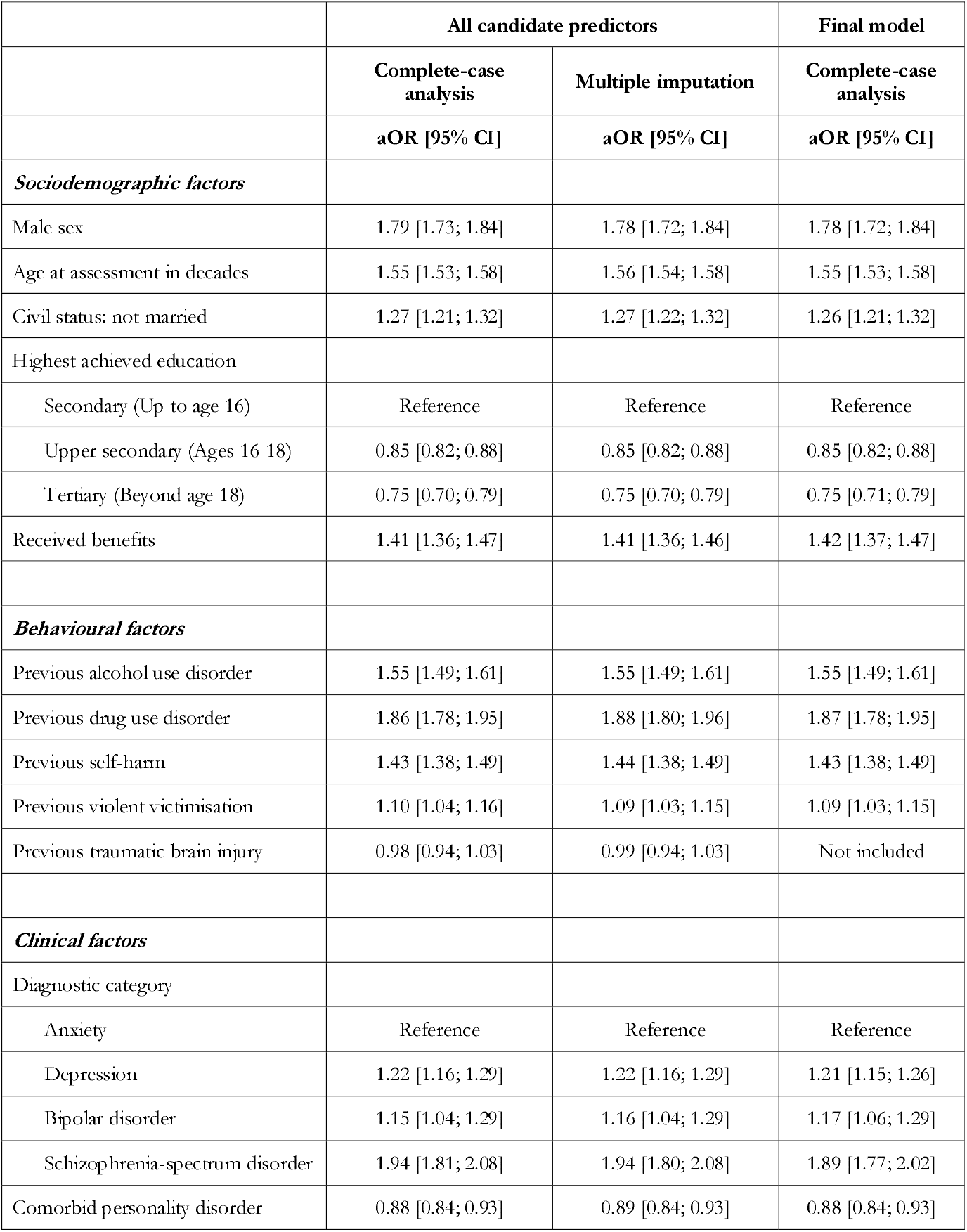
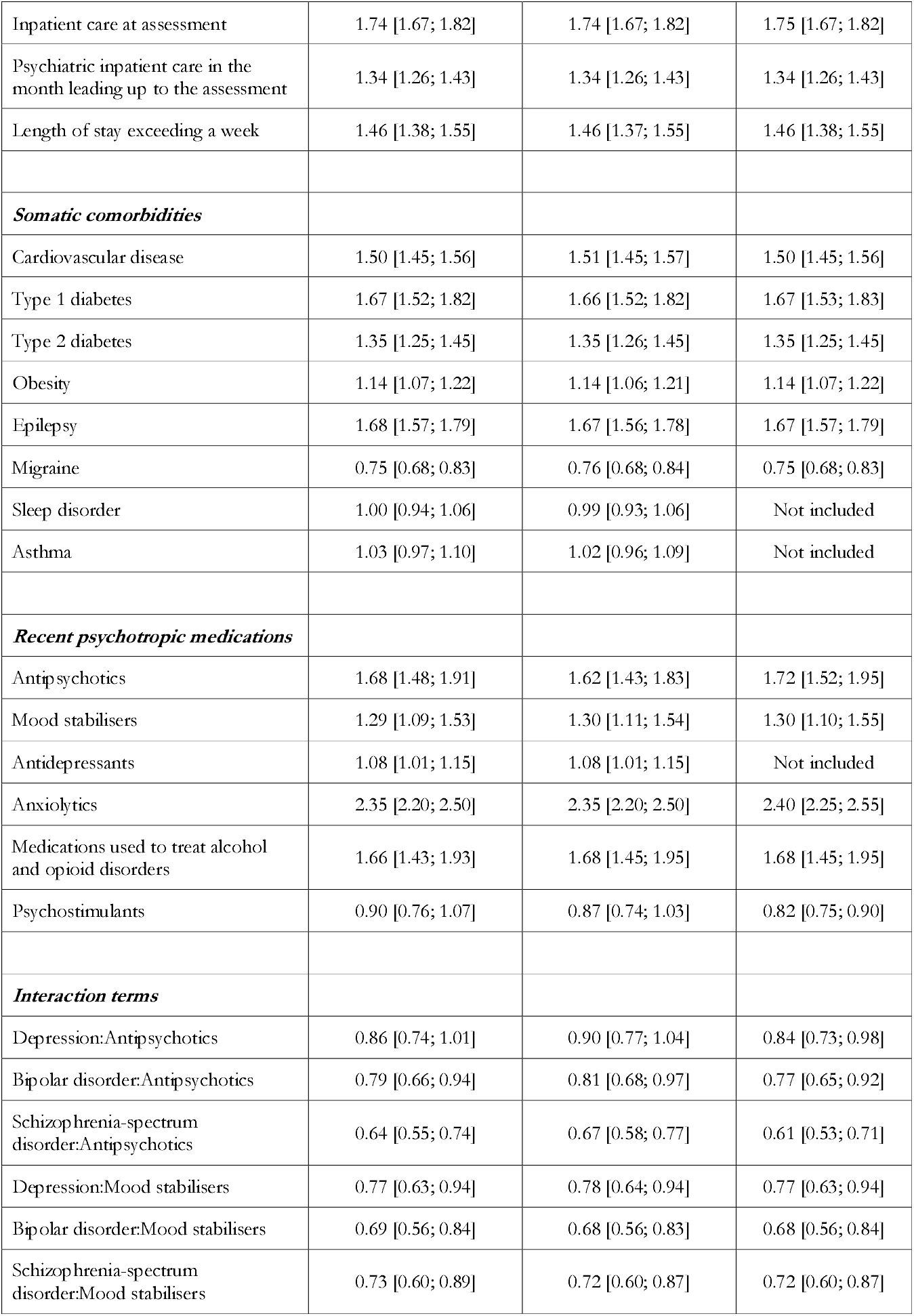
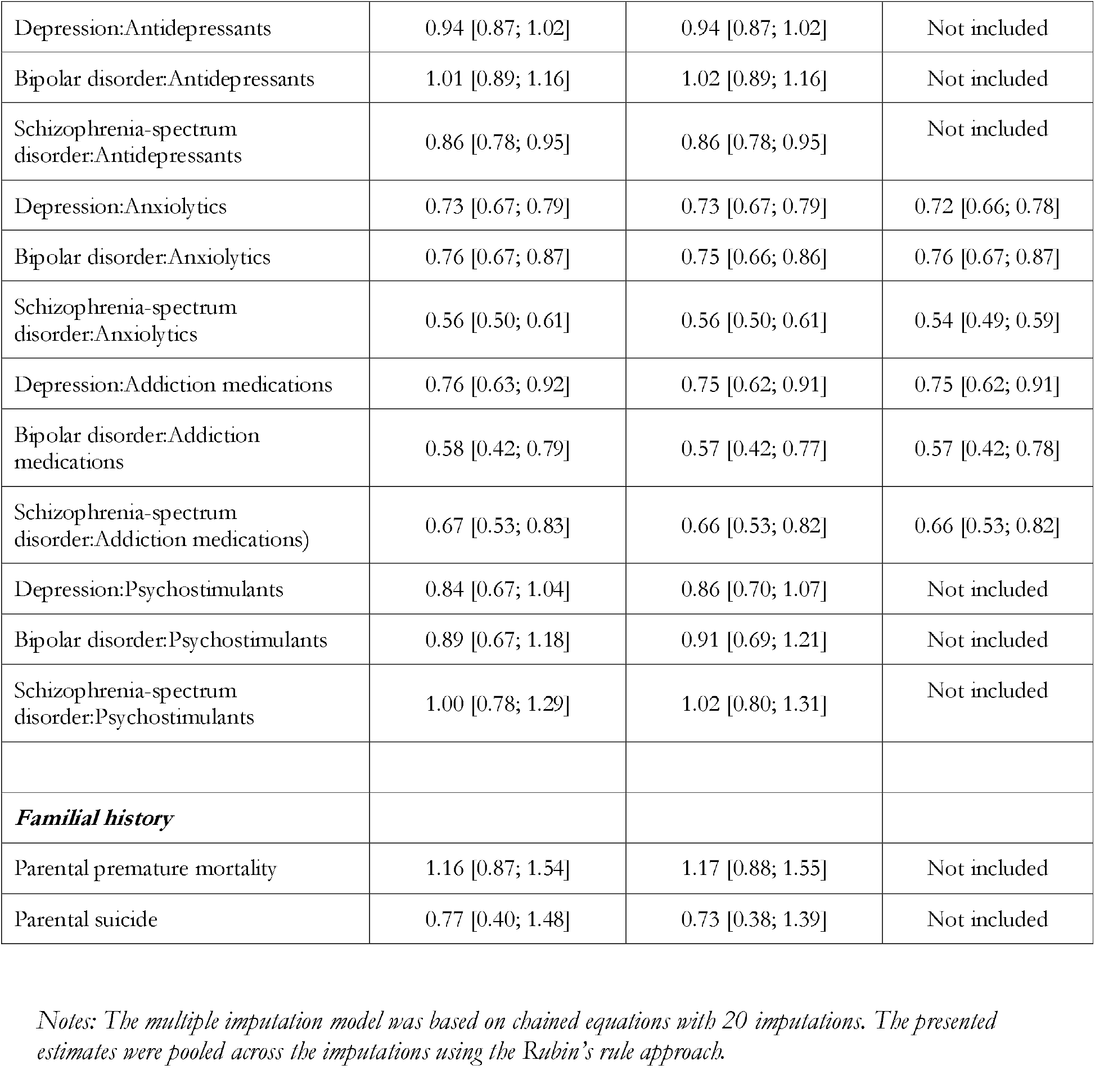
Associations between all candidate predictors (and those retained in the final model) and all-cause mortality within five years of assessment, examined across both complete-case analysis and multiple imputation for missing data.

|  | All candidate predictors |  | Final model |
| --- | --- | --- | --- |
|  | Complete-case analysis | Multiple imputation | Complete-case analysis |
|  | aOR [95% CI] | aOR [95% CI] | aOR [95% CI] |
| ***Sociodemographic factors*** |  |  |  |
| Male sex | 1.79 [1.73; 1.84] | 1.78 [1.72; 1.84] | 1.78 [1.72; 1.84] |
| Age at assessment in decades | 1.55 [1.53; 1.58] | 1.56 [1.54; 1.58] | 1.55 [1.53; 1.58] |
| Civil status: not married | 1.27 [1.21; 1.32] | 1.27 [1.22; 1.32] | 1.26 [1.21; 1.32] |
| Highest achieved education |  |  |  |
| Secondary (Up to age 16) | Reference | Reference | Reference |
| Upper secondary (Ages 16-18) | 0.85 [0.82; 0.88] | 0.85 [0.82; 0.88] | 0.85 [0.82; 0.88] |
| Tertiary (Beyond age 18) | 0.75 [0.70; 0.79] | 0.75 [0.70; 0.79] | 0.75 [0.71; 0.79] |
| Received benefits | 1.41 [1.36; 1.47] | 1.41 [1.36; 1.46] | 1.42 [1.37; 1.47] |
| ***Behavioural factors*** |  |  |  |
| Previous alcohol use disorder | 1.55 [1.49; 1.61] | 1.55 [1.49; 1.61] | 1.55 [1.49; 1.61] |
| Previous drug use disorder | 1.86 [1.78; 1.95] | 1.88 [1.80; 1.96] | 1.87 [1.78; 1.95] |
| Previous self-harm | 1.43 [1.38; 1.49] | 1.44 [1.38; 1.49] | 1.43 [1.38; 1.49] |
| Previous violent victimisation | 1.10 [1.04; 1.16] | 1.09 [1.03; 1.15] | 1.09 [1.03; 1.15] |
| Previous traumatic brain injury | 0.98 [0.94; 1.03] | 0.99 [0.94; 1.03] | Not included |
| ***Clinical factors*** |  |  |  |
| Diagnostic category |  |  |  |
| Anxiety | Reference | Reference | Reference |
| Depression | 1.22 [1.16; 1.29] | 1.22 [1.16; 1.29] | 1.21 [1.15; 1.26] |
| Bipolar disorder | 1.15 [1.04; 1.29] | 1.16 [1.04; 1.29] | 1.17 [1.06; 1.29] |
| Schizophrenia-spectrum disorder | 1.94 [1.81; 2.08] | 1.94 [1.80; 2.08] | 1.89 [1.77; 2.02] |
| Comorbid personality disorder | 0.88 [0.84; 0.93] | 0.89 [0.84; 0.93] | 0.88 [0.84; 0.93] |
| Inpatient care at assessment | 1.74 [1.67; 1.82] | 1.74 [1.67; 1.82] | 1.75 [1.67; 1.82] |
| Psychiatric inpatient care in the month leading up to the assessment | 1.34 [1.26; 1.43] | 1.34 [1.26; 1.43] | 1.34 [1.26; 1.43] |
| Length of stay exceeding a week | 1.46 [1.38; 1.55] | 1.46 [1.37; 1.55] | 1.46 [1.38; 1.55] |
| ***Somatic comorbidities*** |  |  |  |
| Cardiovascular disease | 1.50 [1.45; 1.56] | 1.51 [1.45; 1.57] | 1.50 [1.45; 1.56] |
| Type 1 diabetes | 1.67 [1.52; 1.82] | 1.66 [1.52; 1.82] | 1.67 [1.53; 1.83] |
| Type 2 diabetes | 1.35 [1.25; 1.45] | 1.35 [1.26; 1.45] | 1.35 [1.25; 1.45] |
| Obesity | 1.14 [1.07; 1.22] | 1.14 [1.06; 1.21] | 1.14 [1.07; 1.22] |
| Epilepsy | 1.68 [1.57; 1.79] | 1.67 [1.56; 1.78] | 1.67 [1.57; 1.79] |
| Migraine | 0.75 [0.68; 0.83] | 0.76 [0.68; 0.84] | 0.75 [0.68; 0.83] |
| Sleep disorder | 1.00 [0.94; 1.06] | 0.99 [0.93; 1.06] | Not included |
| Asthma | 1.03 [0.97; 1.10] | 1.02 [0.96; 1.09] | Not included |
| ***Recent psychotropic medications*** |  |  |  |
| Antipsychotics | 1.68 [1.48; 1.91] | 1.62 [1.43; 1.83] | 1.72 [1.52; 1.95] |
| Mood stabilisers | 1.29 [1.09; 1.53] | 1.30 [1.11; 1.54] | 1.30 [1.10; 1.55] |
| Antidepressants | 1.08 [1.01; 1.15] | 1.08 [1.01; 1.15] | Not included |
| Anxiolytics | 2.35 [2.20; 2.50] | 2.35 [2.20; 2.50] | 2.40 [2.25; 2.55] |
| Medications used to treat alcohol and opioid disorders | 1.66 [1.43; 1.93] | 1.68 [1.45; 1.95] | 1.68 [1.45; 1.95] |
| Psychostimulants | 0.90 [0.76; 1.07] | 0.87 [0.74; 1.03] | 0.82 [0.75; 0.90] |
| ***Interaction terms*** |  |  |  |
| Depression:Antipsychotics | 0.86 [0.74; 1.01] | 0.90 [0.77; 1.04] | 0.84 [0.73; 0.98] |
| Bipolar disorder:Antipsychotics | 0.79 [0.66; 0.94] | 0.81 [0.68; 0.97] | 0.77 [0.65; 0.92] |
| Schizophrenia-spectrum disorder:Antipsychotics | 0.64 [0.55; 0.74] | 0.67 [0.58; 0.77] | 0.61 [0.53; 0.71] |
| Depression:Mood stabilisers | 0.77 [0.63; 0.94] | 0.78 [0.64; 0.94] | 0.77 [0.63; 0.94] |
| Bipolar disorder:Mood stabilisers | 0.69 [0.56; 0.84] | 0.68 [0.56; 0.83] | 0.68 [0.56; 0.84] |
| Schizophrenia-spectrum disorder:Mood stabilisers | 0.73 [0.60; 0.89] | 0.72 [0.60; 0.87] | 0.72 [0.60; 0.87] |
| Depression:Antidepressants | 0.94 [0.87; 1.02] | 0.94 [0.87; 1.02] | Not included |
| Bipolar disorder:Antidepressants | 1.01 [0.89; 1.16] | 1.02 [0.89; 1.16] | Not included |
| Schizophrenia-spectrum disorder:Antidepressants | 0.86 [0.78; 0.95] | 0.86 [0.78; 0.95] | Not included |
| Depression:Anxiolytics | 0.73 [0.67; 0.79] | 0.73 [0.67; 0.79] | 0.72 [0.66; 0.78] |
| Bipolar disorder:Anxiolytics | 0.76 [0.67; 0.87] | 0.75 [0.66; 0.86] | 0.76 [0.67; 0.87] |
| Schizophrenia-spectrum disorder:Anxiolytics | 0.56 [0.50; 0.61] | 0.56 [0.50; 0.61] | 0.54 [0.49; 0.59] |
| Depression:Addiction medications | 0.76 [0.63; 0.92] | 0.75 [0.62; 0.91] | 0.75 [0.62; 0.91] |
| Bipolar disorder:Addiction medications | 0.58 [0.42; 0.79] | 0.57 [0.42; 0.77] | 0.57 [0.42; 0.78] |
| Schizophrenia-spectrum disorder:Addiction medications) | 0.67 [0.53; 0.83] | 0.66 [0.53; 0.82] | 0.66 [0.53; 0.82] |
| Depression:Psychostimulants | 0.84 [0.67; 1.04] | 0.86 [0.70; 1.07] | Not included |
| Bipolar disorder:Psychostimulants | 0.89 [0.67; 1.18] | 0.91 [0.69; 1.21] | Not included |
| Schizophrenia-spectrum disorder:Psychostimulants | 1.00 [0.78; 1.29] | 1.02 [0.80; 1.31] | Not included |
| ***Familial history*** |  |  |  |
| Parental premature mortality | 1.16 [0.87; 1.54] | 1.17 [0.88; 1.55] | Not included |
| Parental suicide | 0.77 [0.40; 1.48] | 0.73 [0.38; 1.39] | Not included |

*Notes: The multiple imputation model was based on chained equations with 20 imputations. The presented estimates were pooled across the imputations using the Rubin's rule approach.*

**Table 3.** Distribution of included predictors across the derivation sample and external validation sample.

|  | **Derivation sample** | **External validation sample** |
| --- | --- | --- |
| ***Total*** | 530,201 | 254,691 |
| ***Sociodemographic factors*** |  |  |
| Male sex | 217,426 (41.0%) | 105,873 (41.6%) |
| Age at assessment, years ± SD | 37 ± 13 | 36 ± 14 |
| Civil status: not married | 414,120 (78.1%) | 190,273 (74.7%) |
| Highest achieved education |  |  |
| Up to age 16 | 139,762 (26.4%) | 93,715 (36.8%) |
| Ages 16-18 | 312,654 (59.0%) | 131,166 (51.5%) |
| Beyond age 18 | 77,785 (14.7%) | 29,810 (11.7%) |
| Received benefits | 148,500 (28.0%) | 96,061 (37.7%) |
| ***Behavioural factors*** |  |  |
| Previous alcohol use disorder | 65,140 (12.3%) | 39,331 (15.4%) |
| Previous drug use disorder | 41,243 (7.8%) | 12,844 (5.0%) |
| Previous self-harm | 68,996 (13.0%) | 25,303 (9.9%) |
| Previous violent victimisation | 29,554 (5.6%) | 7862 (3.1%) |
| ***Clinical factors*** |  |  |
| Diagnostic category |  |  |
| Anxiety | 213,878 (40.3%) | 43,816 (17.2%) |
| Depression | 233,368 (44.0%) | 140,767 (55.3%) |
| Bipolar disorder | 36,076 (6.8%) | 19,317 (7.6%) |
| Schizophrenia-spectrum disorder | 46,879 (8.8%) | 50,791 (19.9%) |
| Comorbid personality disorder | 42,295 (8.0%) | 36,127 (14.2%) |
| Inpatient care at assessment | 68,689 (13.0%) | 30,941 (12.1%) |
| Psychiatric inpatient care in the month leading up to the assessment | 14,480 (2.7%) | 8001 (3.1%) |
| Length of stay exceeding a week | 18,829 (3.6%) | 16,285 (6.4%) |
| ***Somatic comorbidities*** |  |  |
| Cardiovascular disease | 55,294 (10.4%) | 30,563 (12.0%) |
| Type 1 diabetes | 9508 (1.8%) | 4254 (1.7%) |
| Type 2 diabetes | 13,914 (2.6%) | 6581 (2.6%) |
| Obesity | 23,849 (4.5%) | 8444 (3.3%) |
| Epilepsy | 13,135 (2.5%) | 8595 (3.4%) |
| Migraine | 17,243 (3.3%) | 10,060 (3.9%) |
| ***Recent psychotropic medications*** |  |  |
| Antipsychotics | 49,888 (9.4%) | 53,658 (21.1%) |
| Mood stabilisers | 33,737 (6.4%) | 16,059 (6.3%) |
| Anxiolytics | 86,529 (16.3%) | 28,820 (11.3%) |
| Medications used to treat alcohol and<br>opioid disorders | 7161 (1.4%) | 731 (0.3%) |
| Psychostimulants | 18,597 (3.5%) | 1220 (0.5%) |

Receiver operating characteristics (ROC) curves are presented in **Figure 1**. The final model demonstrated excellent discriminatory performance in the derivation sample, with an optimism-adjusted area under the ROC curve (AUC) of 0.823. This performance was marginally reduced in the external validation sample (AUC: 0.803; 95% CI: 0.799-0.807).

**Figure 1.**
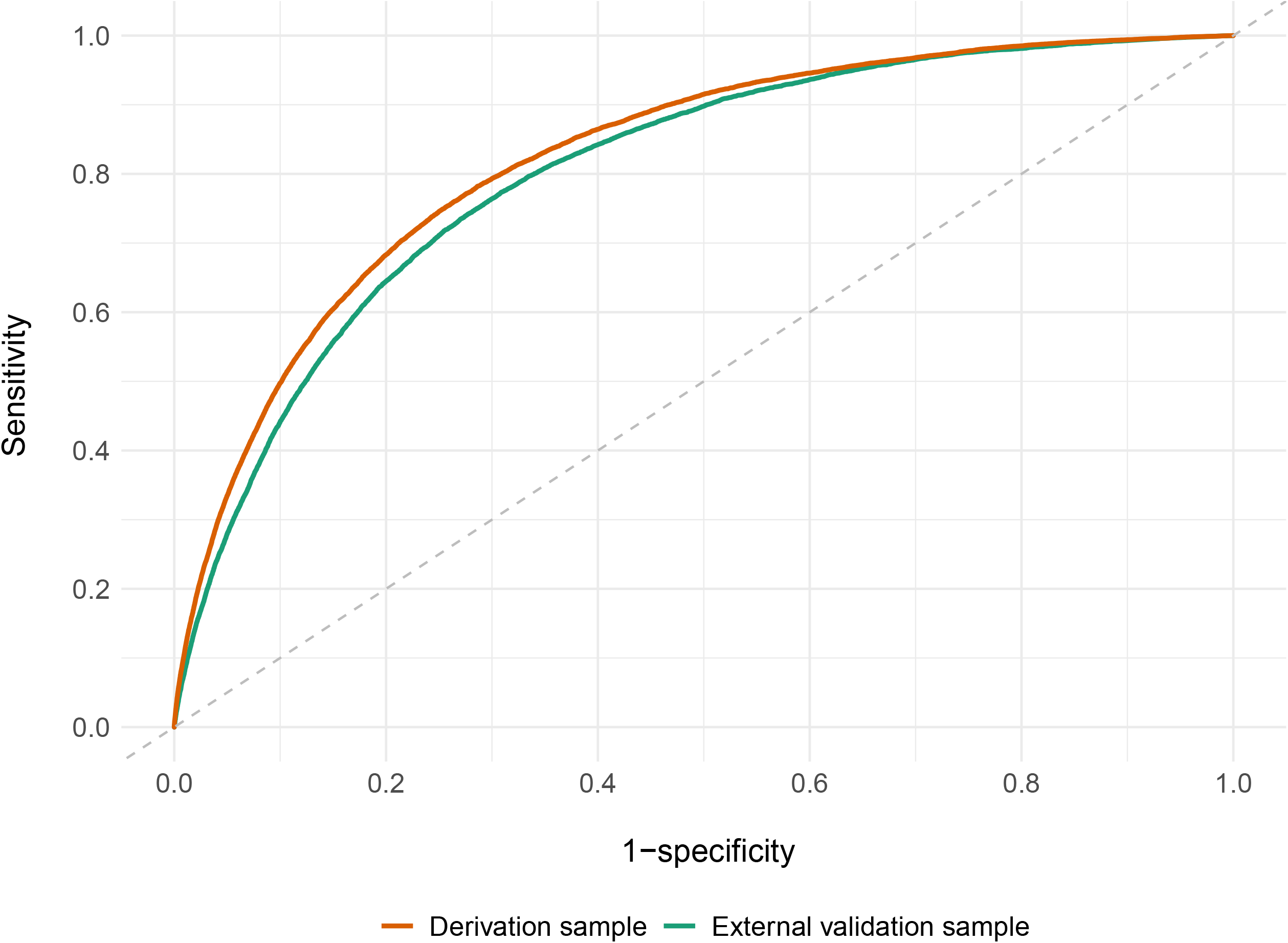
Model discrimination shown by receiver operating characteristics curves for all-cause mortality occurring within 5 years of assessment in the derivation and external validation samples Notes: The optimism-adjusted AUC was 0.823 in the derivation sample, and the equivalent AUC in the external validation was 0.803 (95% CI: 0.799-0.807).

Calibration metrics, adjusted for optimism in the derivation sample, showed strong agreement between predicted and observed risks, with calibration-in-the-large (CITL) close to zero (−0.006) and a calibration slope near 1 (0.998). The calibration plot (**Figure 2**) and associated metrics (ICI: 0.003; E90: 0.011; Emax: 0.259) revealed slight overestimation for predicted probabilities exceeding 20 percent, affecting a small proportion of participants (n=12,234; 2.2%). Predictions from the final model aligned well with those derived from bootstrap resamples (**eFigure 3**), with a median difference within the 95% stability interval of 0.2%. Additionally, pooled estimates from internal-external cross-validation closely matched the optimism-adjusted results (**eFigure 4**). The absolute differences in the performance metrics were negligible across Swedish counties (τ^2^<0.005).

**Figure 2.**
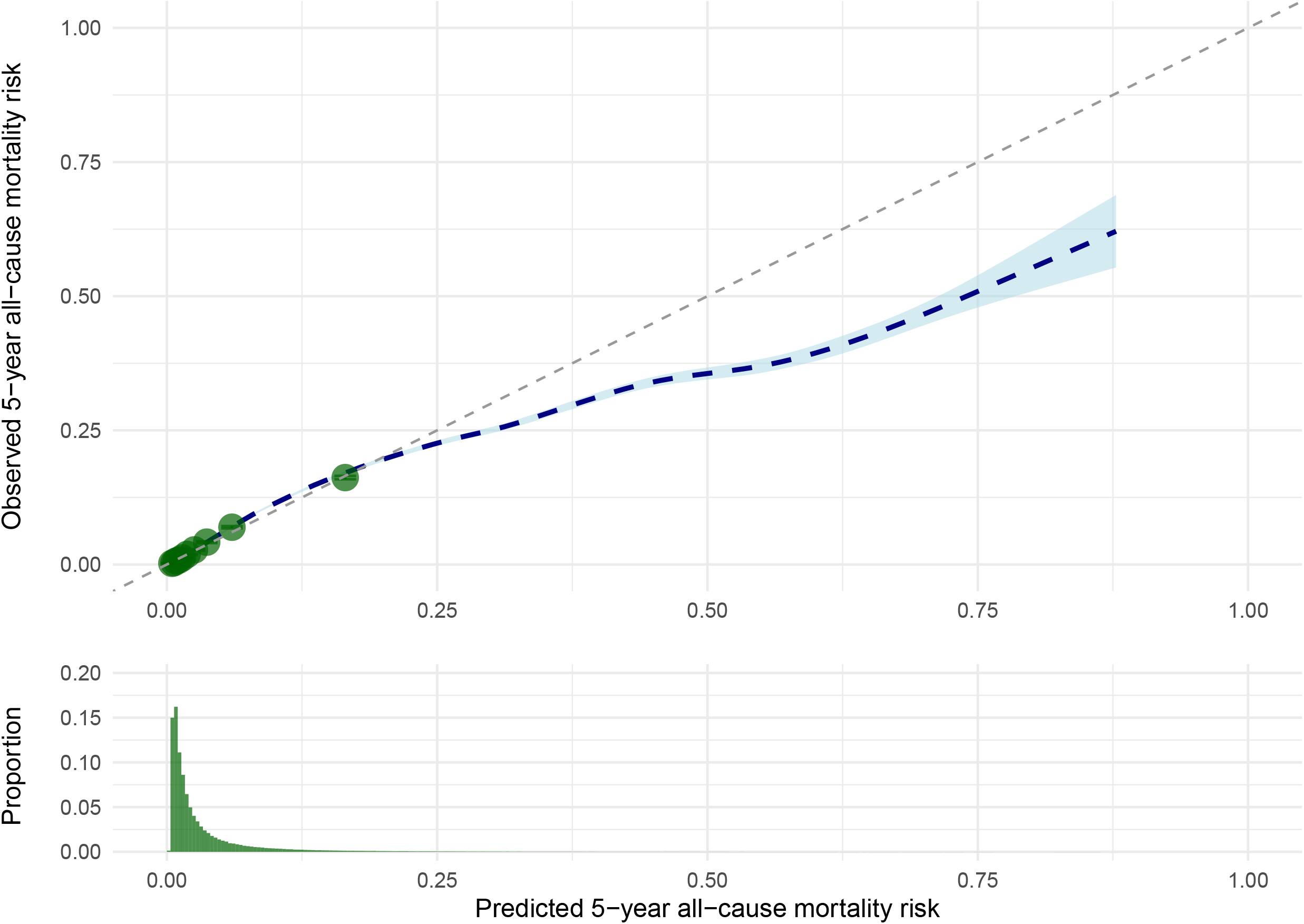
Calibration of the model and distribution of predicted 5-year all-cause mortality risk among individuals with psychiatric disorders in the derivation sample Notes: The green dots represent the average calibration of groups formed by deciles of the predicted risk distribution.

In external validation, the model initially underestimated risks (CITL: 0.47; 95% CI: 0.45 to 0.49), which was expected, given the higher mortality rate and distribution of risk factors in the validation compared with derivation sample (4.4% vs. 3.5%). However, after recalibration of the intercept, calibration closely matched the derivation sample (**Figure 3**; ICI: 0.006; E90: 0.013; Emax: 0.318). In subgroup analyses (**eFigure 5**), the model demonstrated robust performance across all 19 Finnish regions (AUCs: 0.77-0.82). The model performance was slightly higher for women compared with men (AUCs: 0.81 vs. 0.76) and for individuals under 40 years compared with older age categories (AUCs: 0.77-0.78 vs. 0.73-0.75). It was also marginally higher among those who had not received benefits in the year before assessment (AUCs: 0.79 vs. 0.75). Calibration metrics remained excellent in all subgroups, with maximum values for ICI, E90, and Emax of 0.011, 0.032, and 0.525, respectively.

**Figure 3.**
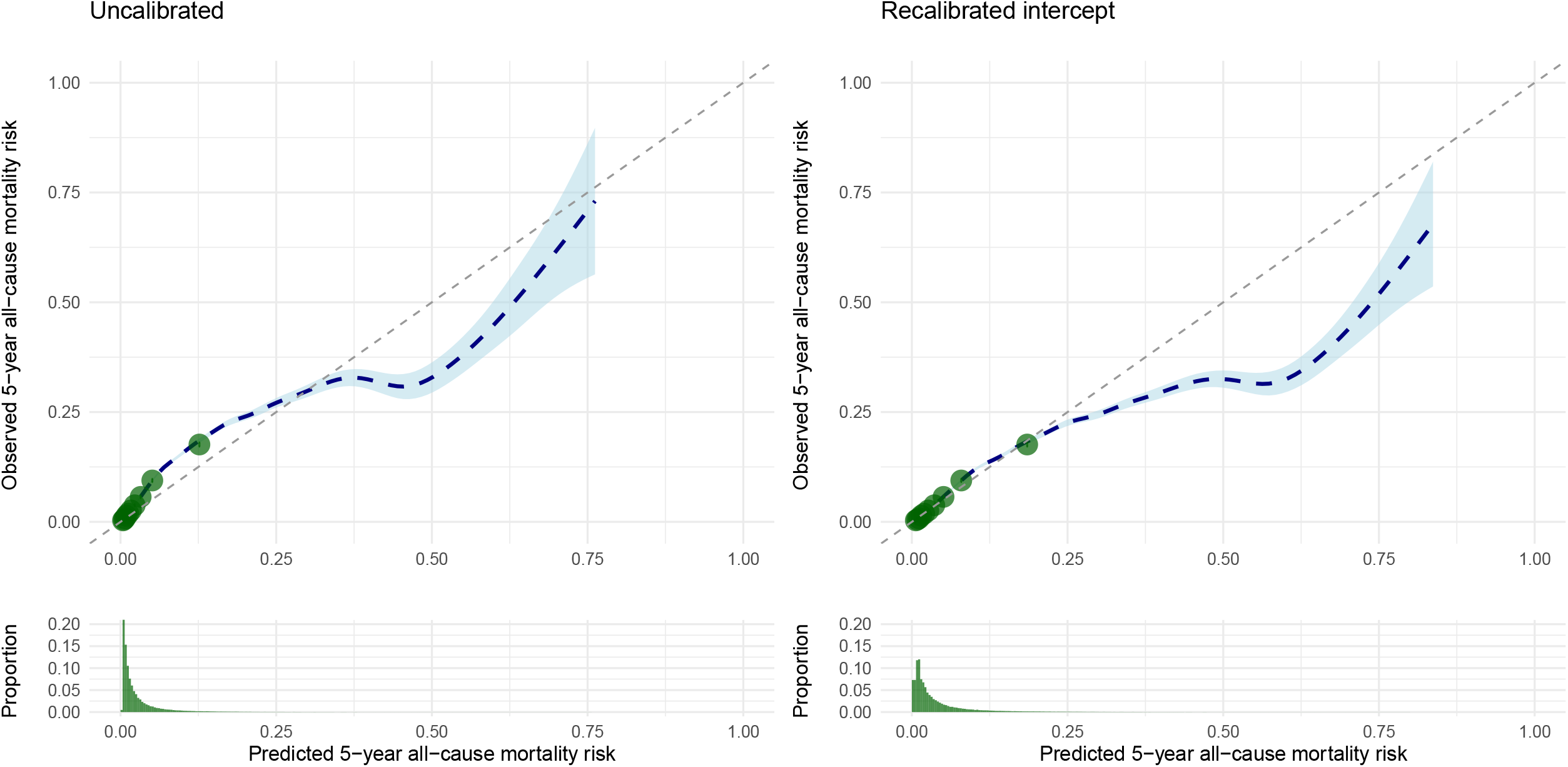
Calibration of the model and distribution of predicted 5-year all-cause mortality risk among individuals with psychiatric disorders in the external validation sample, with and without intercept recalibration Notes: The green dots represent the average calibration of groups formed by deciles of the predicted risk distribution.

The model performed similarly well across all diagnostic categories, although it was somewhat less accurate for individuals with schizophrenia-spectrum disorders (AUCs: 0.78-0.82 vs. 0.73; **eFigure 6**). There were no meaningful differences in model performance when shorter follow-up periods were considered (AUCs: 0.79-0.80). It predicted deaths due to natural causes more accurately than those due to external causes (AUCs: 0.83 vs. 0.75). This difference was largely driven by poorer performance in suicide prediction (AUC: 0.70).

In a post-hoc sensitivity analysis, all individuals in the external validation sample with a predicted mortality risk exceeding 20% (n=7599 or 3.0%) were reassigned a predicted mortality risk of 20%. This ceiling adjustment reduced the maximum difference between observed and predicted mortality risks (Emax) from 31.8% to 3.1%, demonstrating improved calibration (ICI: 0.005; E90: 0.015; Emax: 0.031).

## Discussion

Using data from two nationally representative register-based cohorts comprising nearly 785,000 individuals diagnosed with mental illness, we developed and externally validated a prognostic model to assess risk of all-cause mortality within five years of any secondary care assessment. During follow-up, approximately one in 26 individuals died, with fewer than a third of these being determined as suicides. The model, which integrates 25 routinely collected sociodemographic, and clinical factors, showed excellent discriminatory performance and calibration. In external validation, it achieved an area under the curve (AUC) of 0.803 (95% CI: 0.803-0.803), which means that if a pair of individuals were randomly selected, one who died and one who survived within 5 years of assessment, the model would correctly assign a higher risk to the individual who died in at least 80 percent of cases. The Integrated Calibration Index indicated that the average difference between predicted and observed mortality risks was 0.6%.

The model has been translated into a freely available web-based calculator, providing a five-year mortality risk score for each individual. As it is based on routinely collected data and predictors that are easily scored, it offers a feasible tool for research, training and clinical use. Although overall calibration demonstrated a close alignment between predicted and observed risks, the model overpredicted risk among very high-risk individuals whose predicted risk exceeded 20%, which constituted no more than 3% of the samples. Sensitivity analyses revealed improved calibration when a 20% ceiling was imposed on predicted risk. Consequently, we set a maximum of 20% predicted risk in the calculator to mitigate spurious precision for individuals at very high risk. This modification reflects the likelihood that important contextual risk factors relevant to high-risk individuals are not fully captured by routinely collected data, and it underscores the need for clinical judgement to augment the use of the tool in practice.

To our knowledge, the only other risk stratification tool specifically designed to predict all-cause mortality in individuals with mental illness is the MIRACLE-FEP tool,^28^ although its use is limited to those with first-episode psychosis. Its discriminatory accuracy in external validation (AUC range: 0.67-0.70) was lower than that observed in the present study, but these estimates were characterised by considerable uncertainty due to small external validation samples, with the smallest comprising 1490 participants. Furthermore, the calibration of the tool across the full spectrum of predicted risks remains unknown. Contrary to expert guidelines,^29^ calibration plots for this tool used varying numbers of binned groups across samples and were restricted to a 5-6% predicted mortality risk, thereby leaving broader calibration across the entire range of predicted risks unexplored. A further external validation in first-episode bipolar disorder showed similar performance.^30^

An important consideration prior to implementing the tool in clinical care settings is understanding the extent to which patients want to know their risk of premature mortality. In our focus group meetings with individuals with lived experience of mental illness, the overwhelming majority expressed a strong desire to be informed of their premature mortality risk level. At the same time, they emphasised the importance of the information being delivered in a sensitive manner by their clinicians, along with practical, personalised advice on strategies to reduce their risk.

The use of mortality risk prognostic tools will not impact mortality rates unless they are linked with effective interventions targeting modifiable risk factors for premature mortality. Evidence from a representative UK-based study^31^ suggested that only about half (55%) of patients with severe mental illnesses had been comprehensively screened for cardiometabolic risk factors at least once between 2000 and 2018. Ideally, all individuals with mental illness should receive screening of cardiometabolic risk factors, including substance misuse, on an annual basis, and more frequently for those at elevated predicted risk of 5-year mortality. This approach could potentially reduce risks of either undiagnosed or late-diagnosed physical diseases affecting patients with mental illness, both of which contribute to poorer prognoses.^5,32^ Risk assessment tools for cardiovascular disease, such as QR4,^33^ have typically been developed using primary care registers, and tend to perform less well in secondary care settings.^34^ Importantly, approximately half of all deaths were in the present study attributed to unnatural causes, and there was large heterogeneity in the underlying causes of natural deaths across mental illnesses. For these reasons, it is likely that MortOx can provide more accurate risk predictions than other models. In fact, our sensitivity analyses demonstrated that MortOx performed similarly well in predicting cause-specific mortality, though somewhat poorer for suicide, where its performance was nevertheless similar to that of existing externally validated suicide risk prediction tools.^35^

Whilst screening remains an important tool for the early detection of potential physical health problems, other types of multiprofessional preventive interventions are needed to motivate patients to make substantial lifestyle changes (e.g. physical activity, nutrition, reducing smoking, and limiting alcohol). There is currently mixed support for such interventions in individuals with mental illness, but an expert review recently suggested that developing higher-quality interventions, which encompass a wider range of professions (e.g. dietitians and exercise professionals) and improved access to supervised exercise services, may lead to better outcomes.^1^ This highlights the need to implement a more integrated approach to care, focusing on preventing and treating both mental and physical health outcomes,^36^ particularly in those at elevated predicted risk of 5-year mortality.

Key strengths of this study include the use of large-scale, nationally representative datasets with minimal selection bias, enabling the development of a model to be based on a cohort of 530,201 individuals, among whom 18,619 deaths occurred within five years of assessment. By utilising a multivariate modelling approach, we found that many previously identified risk factors (e.g. traumatic brain injury and parental premature mortality) were not independently associated with increased premature mortality risk. A novel feature of the model was the inclusion of multiple interaction terms, which allowed the associations between recent psychotropic drug use and mortality risk to vary across diagnostic categories. This underscores the complexity of these associations and highlights the limitations of clinical judgement in capturing such nuances. Furthermore, we employed rigorous internal validation techniques, including performance evaluation through bootstrapping, which replicated the full analytic procedures of model development, and allowed us to generate a stability plot to assess the stability of individual predictions from the model. The large sample size further allowed for assessment of geographical differences in model performance through an internal-external validation approach. Additionally, the model was evaluated on an independent cohort of 254,691 Finnish patients, among whom 11,206 deaths were recorded during follow-up. The latter cohort was sufficiently large to allow for detailed subgroup analyses to assess the algorithmic fairness of the model.

Some limitations should be noted. First, although the use of routinely collected register data enabled us to analyse large cohorts of individuals with mental illness, they do not capture all potentially relevant predictors, such as psychiatric symptoms, medication non-adherence, and lifestyle factors (e.g., smoking, physical activity, and nutrition). The extent to which incorporating these additional factors might incrementally improve the model, beyond the 25 included predictors, could be examined in future research. Second, although the model performed well across Swedish counties, the external validation in Finland highlighted that large differences in country-specific mortality rates may require a recalibration of the model intercept. Third, the model development relied on complete-case analyses, which excluded a small proportion of missing data (5%) from the derivation cohort. However, sensitivity analyses conducted using multiple imputation techniques revealed negligible differences in results. This, combined with the findings from the model stability analyses, suggests that our model was robust to alternative model specifications and variations in the underlying populations. Fourth, outpatient care visit data were unavailable prior to 2001 in Sweden and 1998 in Finland. This limitation implies that many predictors related to earlier behavioural problems and comorbidities may primarily reflect more severe forms of these conditions during those periods.

## Conclusions

In summary, we have developed and validated a transdiagnostic clinical prognostic tool that estimates 5-year mortality risk in individuals with mental illness. How this model can improve outcomes needs research on implementation, linkage to effective intervention, and net benefit compared with current practice.

## Data Availability

Due to Swedish and Finnish privacy legislation, individual-level data cannot be shared publicly. Researchers wishing to replicate this work can apply for data access through Statistics Sweden (https://www.scb.se/en/services/ordering-data-and-statistics/ordering-microdata/), the Swedish National Board of Health and Welfare (https://www.socialstyrelsen.se/en/statistics-and-data/registers/), and Findata (https://findata.fi/en/).

## Author contributions

AS and SF contributed to the conception and design of the study. PL obtained linked data from national Swedish registers, and PM from national Finnish registers. AS conducted the statistical analyses and drafted the paper with SF. All authors interpreted the findings, critically revised the draft, approved the final version of the manuscript, and take full responsibility for the accuracy and integrity of the work.

## References

1. Firth J, Siddiqi N, Koyanagi A, et al. The Lancet Psychiatry Commission: a blueprint for protecting physical health in people with mental illness. The Lancet Psychiatry. 2019;6(8):675–712.

2. Chesney E, Goodwin GM, Fazel S. Risks of all-cause and suicide mortality in mental disorders: a meta-review. World Psychiatry. 2014;13(2):153–160.

3. Chan JKN, Correll CU, Wong CSM, et al. Life expectancy and years of potential life lost in people with mental disorders: a systematic review and meta-analysis. eClinicalMedicine. 2023;65:102294.

4. Momen NC, Plana-Ripoll O, Agerbo E, et al. Mortality associated with mental disorders and comorbid general medical conditions. JAMA Psychiatry. 2022;79(5):444–453.

5. Sariaslan A, Sharpe M, Larsson H, Wolf A, Lichtenstein P, Fazel S. Psychiatric comorbidity and risk of premature mortality and suicide among those with chronic respiratory diseases, cardiovascular diseases, and diabetes in Sweden: A nationwide matched cohort study of over 1 million patients and their unaffected siblings. PLoS Med. 2022;19(1):e1003864.

6. Ward A, Ishak K, Proskorovsky I, Caro J. Compliance with refilling prescriptions for atypical antipsychotic agents and its association with the risks for hospitalization, suicide, and death in patients with schizophrenia in Quebec and Saskatchewan: a retrospective database study. Clin Ther. 2006;28(11):1912–1921.

7. Ganna A, Ingelsson E. 5 year mortality predictors in 498 103 UK Biobank participants: A prospective population-based study. Lancet. 2015;386(9993):533–540.

8. Hydoub YM, Walker AP, Kirchoff RW, et al. Risk prediction models for hospital mortality in general medical patients: A systematic review. American Journal of Medicine Open. 2023;10:100044.

9. Goldstein BA, Navar AM, Pencina MJ, Ioannidis JPA. Opportunities and challenges in developing risk prediction models with electronic health records data: a systematic review. J Am Med Inform Assoc. 2017;24(1):198–208.

10. Meehan AJ, Lewis SJ, Fazel S, et al. Clinical prediction models in psychiatry: a systematic review of two decades of progress and challenges. Mol Psychiatry. 2022;27(6):2700–2708.

11. Hiroeh U, Appleby L, Mortensen PB, Dunn G. Death by homicide, suicide, and other unnatural causes in people with mental illness: a population-based study. The Lancet. 2001;358(9299):2110–2112.

12. Walker ER, McGee RE, Druss BG. Mortality in mental disorders and global disease burden implications: A systematic review and meta-analysis. JAMA Psychiatry. 2015;72(4):334–341.

13. Erlangsen A, Andersen PK, Toender A, Laursen TM, Nordentoft M, Canudas-Romo V. Cause-specific life-years lost in people with mental disorders: a nationwide, register-based cohort study. The Lancet Psychiatry. 2017;4(12):937–945.

14. Lawrence D, Kisely S, Pais J. The epidemiology of excess mortality in people with mental illness. Can J Psychiatry. 2010;55(12):752–760.

15. Hayes JF, Marston L, Walters K, King MB, Osborn DPJ. Mortality gap for people with bipolar disorder and schizophrenia: UK-based cohort study 2000–2014. The British Journal of Psychiatry. 2017;211(3):175–181.

16. Chen D, Momen NC, Ejlskov L, et al. Socioeconomic inequalities in mortality associated with mental disorders: a population-based cohort study. World Psychiatry. 2025;24(1):92–102.

17. Ludvigsson J, Nørgaard M, Weiderpass E, et al. Ethical aspects of registry-based research in the Nordic countries. Clinical Epidemiology. 2015;7:491.

18. Collins GS, Moons KGM, Dhiman P, et al. TRIPOD+AI statement: updated guidance for reporting clinical prediction models that use regression or machine learning methods. BMJ. 2024;385:e078378.

19. Ludvigsson J, Almqvist C, Edstedt Bonamy AK, et al. Registers of the Swedish total population and their use in medical research. European Journal of Epidemiology. 2016;31(2):125–136.

20. The Finnish Institute for Health and Welfare (THL). Care Register for Health Care. Accessed July 16, 2022. https://thl.fi/en/web/thlfi-en/statistics-and-data/data-and-services/register-descriptions/care-register-for-health-care

21. Official Statistics of Finland (OSF). Quality Description: Causes of death 2017. Published online 2017. Accessed May 11, 2020. http://www.stat.fi/til/ksyyt/2017/ksyyt_2017_2018-12-17_laa_001_en.html

22. The Swedish National Board for Health and Welfare (Socialstyrelsen). The Statistical Register’s Production and Quality: The Cause of Death Register [Det Statistiska Registrets Framställning Och Kvalitet: Dödsorsaksregistret].; 2022.

23. Momen NC, Plana-Ripoll O, Agerbo E, et al. Association between mental disorders and subsequent medical conditions. The New England journal of medicine. 2020;382(18):1721–1731.

24. Fazel S, Wolf A, Larsson H, Mallett S, Fanshawe TR. The prediction of suicide in severe mental illness: development and validation of a clinical prediction rule (OxMIS). Transl Psychiatry. 2019;9(1):98.

25. Steyerberg EW, Harrell FE. Prediction models need appropriate internal, internal–external, and external validation. Journal of Clinical Epidemiology. 2016;69:245–247.

26. Riley RD, Collins GS. Stability of clinical prediction models developed using statistical or machine learning methods. Biometrical Journal. 2023;65(8):2200302.

27. Van Calster B, McLernon DJ, van Smeden M, Wynants L, Steyerberg EW. Calibration: the Achilles heel of predictive analytics. BMC Medicine. 2019;17(1):1–7.

28. Lieslehto J, Tiihonen J, Lähteenvuo M, et al. Development and validation of a machine learning–based model of mortality risk in first-episode psychosis. JAMA Network Open. 2024;7(3):e240640.

29. Riley RD, Archer L, Snell KIE, et al. Evaluation of clinical prediction models (part 2): how to undertake an external validation study. BMJ. 2024;384:e074820.

30. Lieslehto J, Tiihonen J, Lähteenvuo M, et al. Machine learning-based mortality risk assessment in first-episode bipolar disorder: a transdiagnostic external validation study. eClinicalMedicine. 2025;81:103108.

31. Launders N, Jackson CA, Hayes JF, et al. Prevalence and patient characteristics associated with cardiovascular disease risk factor screening in UK primary care for people with severe mental illness: an electronic healthcare record study. BMJ Ment Health. 2025;28(1):e301409.

32. Liberati E, Kelly S, Price A, et al. Diagnostic inequalities relating to physical healthcare among people with mental health conditions: a systematic review. eClinicalMedicine. 2025;80:103026.

33. Hippisley-Cox J, Coupland CAC, Bafadhel M, et al. Development and validation of a new algorithm for improved cardiovascular risk prediction. Nat Med. 2024;30(5):1440–1447.

34. Parsons RE, Liu X, Collister JA, Clifton DA, Cairns BJ, Clifton L. Independent external validation of the QRISK3 cardiovascular disease risk prediction model using UK Biobank. Heart. 2023;109(22):1690–1697.

35. Sariaslan A, Fanshawe T, Pitkänen J, Cipriani A, Martikainen P, Fazel S. Predicting suicide risk in 137,112 people with severe mental illness in Finland: external validation of the Oxford Mental Illness and Suicide tool (OxMIS). Transl Psychiatry. 2023;13(1):126.

36. Sharpe M, Naylor C. Integration of mental and physical health care: from aspiration to practice. Lancet Psychiatry. 2016;3(4):312–313.

