## Supplementary Materials for "Prediction of 5-year mortality risk in 784,892 people with mental illness: Development and validation of a novel clinical prognostic model (MortOx)"

**eText 1. Minimum sample sizes**

We followed previous publications^1,2^ to calculate the minimum required sample sizes for both the development and external validation datasets. For the development dataset, we tested a series of plausible assumptions regarding the magnitude of the discriminatory accuracy, the number of model parameters, and prevalence rates, which resulted in a minimum required sample size of 17,606 individuals and 881 mortality events. Once we generated the final model, we used its distribution of predicted risks, and model performance metrics, as well as an assumed 5% prevalence rate to calculate a minimum required external validation sample size of 14,819 individuals and 741 mortality events.

Our development dataset (n=530,201; 18,619 deaths) and external validation dataset (n=254,691; 11,206 deaths) substantially exceeded these minimum thresholds, providing robust statistical power for model development and validation.

**References**

1.	Riley, R. D. *et al.* Calculating the sample size required for developing a clinical prediction model. *BMJ* **368**, m441 (2020).

2.	Riley, R. D. *et al.* Minimum sample size for external validation of a clinical prediction model with a binary outcome. *Stat. Med.* **40**, 4230–4251 (2021).

**eText 2. Quotes from the focus group interviews**

***Participant F1***: “I think I would want to know […] but it depends on how you told me. Like, it is so sensitive, isn’t it? Do I really want to know? And obviously you have to give people hope. You can’t just say, ‘Oh, it is quite likely you might die in five years.’ It is more about what can I do now that might help me on that journey. Like, how can I improve those statistics and defy that prediction?”

***Participant F2***: “I’m type 2 diabetic, have depressive bipolar. I have spinal challenges. Obese. You name it. Asthma, smoker. I'm not on any non-medical drugs. I know all this because my doctor very carefully spoke to me about it. She was great. I sat and thought about it.

What has really helped me has been a change in mindset. In terms of my thinking, she has been very good at helping me with that. She doesn't have to, but she does. It's about changing my thinking patterns. Looking at those, using more words of affirmation. I kind of knew it anyway when we had this conversation and got all the blood tests. I don't know if that’s helpful, but I know it has certainly helped me. I make an effort every single day, every day, to make sure that I can at least last a little bit longer than five years. Well, four years now.”

***Participant M1***: “I think definitely signposting is the way. […] I would be looking for the right support, in the right area. I think the emphasis is on the doctors to be really, really sensitive and careful with that type of information. You know, especially if the person already has that feeling of ‘I already know, I just need a concrete yes, it looks like this is the way it is going.’ But then that positive take on what ***Participant F2*** said, what would you like to do, because you might find it slightly reverses itself. It might be more than five years. That is a massive boost.”

***Participant M2***: “I think for me it would be based around quality of life. I think I would like to know, and also to draw up an action plan, which could potentially include psychological interventions, like visiting a health psychologist every month. I think a health psychologist could really help put things into perspective. What would be the best action plan for me to live the best life I can, especially if I had a short time or a shorter time to live. I think that is really, really important.”

***Participant F3***: “Yeah, I want to say that I probably wouldn't want to know. I would want my GP and any other professionals to know. But for me, I think I would worry that I would dwell on it. You know, I’ve already got a prognosis for a rare physical illness I have. I was diagnosed 20 years ago, and the prognosis was anywhere between one and thirty years. So, I’ve kind of lived with that uncertainty, and I’m still living with it.”

**eTable 1. ICD codes**

|  | **ICD-8** | **ICD-9** | **ICD-10** |
| --- | --- | --- | --- |
| **Anxiety** |  |  |  |
| Finland | 300 except 300.4 | 300 except 300.4A | F40-F42, F44-F45, F48 |
| Sweden | 300 except 300.4 | 300 except 300E | F40-F42, F44-F45, F48 |
| **Depression** |  |  |  |
| Finland | 296.2, 300.4 | 296.1 [excl. 296.1E],  296.8A, 300.4A | F32-F39  [excl. F32.3 and F33.3] |
| Sweden | 296.2, 300.4 | 296B, 300E, 311 | F32-F39  [excl. F32.3 and F33.3] |
| **Bipolar disorder** |  |  |  |
| Finland | 296  [excl. 296.2] | 296  [excl. 296.1A-D, 296.1F-G] | F30-F31 |
| Sweden | 296  [excl. 296.2] | 296  [excl. 296B] | F30-F31 |
| **Schizophrenia-spectrum disorder** | 295, 297-299 | 295, 297-298 | F20-F29 |
| **Personality disorder** | 301 | 301 | F60-F69 |
| **Alcohol use disorder** |  |  |  |
| Finland | 291, 303 | 291, 303, 305.0 | F10 [excl. F10.5] |
| Sweden | 291, 303 | 291, 303, 305A | F10 [excl. F10.5] |
| **Drug use disorder** |  |  |  |
| Finland | 304 | 292, 304, 305.2-305.9 | F11-F12, F14-F16, F19  [excl. F1*.5] |
| Sweden  Sweden | 304 | 292, 304, 305X | F11-F12, F14-F16, F19  [excl. F1*.5] |
| **Violent victimisation** | E960-E969 | E960-E969 | X85-X99, Y00-Y09 |
| **Self-harm** |  |  |  |
| Finland | - | E950A-E959X,  E970A-E979A | X60-X84,  Y10-Y34 |
| Sweden | E950-E959,  E980-E989 | E950-E959,  E980-E989 | X60-X84,  Y10-Y34 |
| **Cardiovascular disease** | 390-438, 440, 444, 445,  450-453, 458 | 390-438, 440, 444 | I00-I70, I73.0, I74-I75 |
| **Type 1 diabetes** | - | - | E10 |
| **Type 2 diabetes** | - | - | E11 |
| **Obesity** |  |  |  |
| Finland | 277 | 2780A, 2781A | E65-E66 |
| Sweden | 277 | 278A, 278B | E65-E66 |
| **Epilepsy** | 345 | 345 | G40-G41 |
| **Migraine** |  |  |  |
| Finland | 346.09 | 3460A, 3461A, 3469X | G43 |
| Sweden | 346.09 | 346A, 346B, 346W, 346X | G43 |
| **Cause-specific mortality** |  |  |  |
| External causes | - | - | V00-V99, W00-W99,  X00-X99, Y00-Y98 |
| Accidents | - | - | V00-V99, W00-W99,  Y00-Y59 |
| Substance use disorders | - | - | F10-F12, F14-F16, F19  [excl. F1*.5] |
| **Suicide** | - | - | X60-X84,  Y10-Y34 |

*Notes: Deaths from natural causes are defined as those occurring in the absence of any external causes.*

**eTable 2. Final model coefficients and their associated adjusted odds ratios (aORs) with**

**95% confidence intervals (CIs)**

|  | **Coefficient** | **aOR [95% CI]** |
| --- | --- | --- |
| Intercept | -6.485 | 0.00 [0.00; 0.00] |
| Male sex | 0.577 | 1.78 [1.72; 1.84] |
| Age at assessment (in decades) | 0.440 | 1.55 [1.53; 1.58] |
| Civil status: not married | 0.235 | 1.26 [1.21; 1.32] |
| Educational attainment: Up to age 16 | Reference |  |
| Educational attainment: Ages 16-18 | -0.161 | 0.85 [0.82; 0.88] |
| Educational attainment: Beyond age 18 | -0.290 | 0.75 [0.71; 0.79] |
| Received benefits | 0.347 | 1.42 [1.37; 1.47] |
| Previous alcohol use disorder | 0.438 | 1.55 [1.49; 1.61] |
| Previous drug use disorder | 0.623 | 1.87 [1.78; 1.95] |
| Previous self-harm | 0.358 | 1.43 [1.38; 1.49] |
| Previous violent victimisation | 0.087 | 1.09 [1.03; 1.15] |
| Comorbid personality disorder | -0.128 | 0.88 [0.84; 0.93] |
| Inpatient care at assessment | 0.557 | 1.75 [1.67; 1.82] |
| Length of stay exceeding a week | 0.379 | 1.46 [1.38; 1.55] |
| Psychiatric inpatient care in the month leading up to the assessment | 0.296 | 1.34 [1.26; 1.43] |
| Cardiovascular disease | 0.408 | 1.50 [1.45; 1.56] |
| Type 1 diabetes | 0.512 | 1.67 [1.53; 1.83] |
| Type 2 diabetes | 0.298 | 1.35 [1.25; 1.45] |
| Obesity | 0.134 | 1.14 [1.07; 1.22] |
| Epilepsy | 0.515 | 1.67 [1.57; 1.79] |
| Migraine | -0.283 | 0.75 [0.68; 0.83] |
| Diagnostic category: Anxiety | Reference |  |
| Diagnostic category: Depression | 0.187 | 1.21 [1.15; 1.26] |
| Diagnostic category: Bipolar disorder | 0.157 | 1.17 [1.06; 1.29] |
| Diagnostic category: Schizophrenia-spectrum disorder | 0.637 | 1.89 [1.77; 2.02] |
| Antipsychotics | 0.544 | 1.72 [1.52; 1.95] |
| Mood stabilisers | 0.265 | 1.30 [1.10; 1.55] |
| Anxiolytics | 0.875 | 2.40 [2.25; 2.55] |
| Medications used to treat alcohol and opioid disorders | 0.517 | 1.68 [1.45; 1.95] |
| Psychostimulants | -0.196 | 0.82 [0.75; 0.90] |
| *Interaction terms* |  |  |
| Depression:Antipsychotics | -0.170 | 0.84 [0.73; 0.98] |
| Bipolar disorder:Antipsychotics | -0.256 | 0.77 [0.65; 0.92] |
| Schizophrenia-spectrum disorder:Antipsychotics | -0.488 | 0.61 [0.53; 0.71] |
| Depression:Mood stabilisers | -0.264 | 0.77 [0.63; 0.94] |
| Bipolar disorder:Mood stabilisers | -0.380 | 0.68 [0.56; 0.84] |
| Schizophrenia-spectrum disorder:Mood stabilisers | -0.325 | 0.72 [0.60; 0.87] |
| Depression:Anxiolytics | -0.335 | 0.72 [0.66; 0.78] |
| Bipolar disorder:Anxiolytics | -0.271 | 0.76 [0.67; 0.87] |
| Schizophrenia-spectrum disorder:Anxiolytics | -0.622 | 0.54 [0.49; 0.59] |
| Depression:Addiction medications | -0.286 | 0.75 [0.62; 0.91] |
| Bipolar disorder:Addiction medications | -0.556 | 0.57 [0.42; 0.78] |
| Schizophrenia-spectrum disorder:Addiction medications) | -0.418 | 0.66 [0.53; 0.82] |

**eFigure 1. Sample flowcharts**

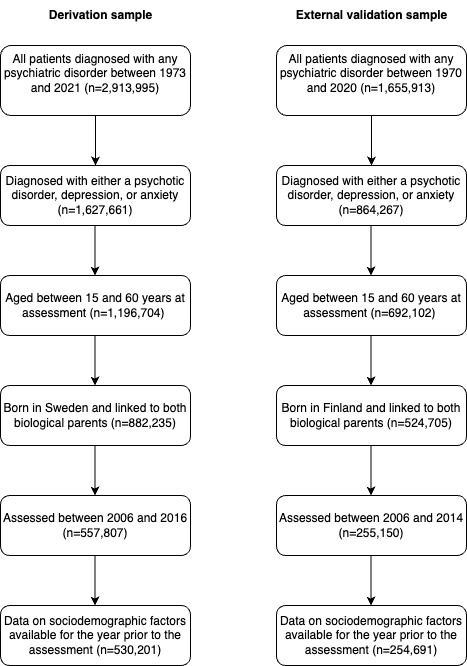

**eFigure 2. Percentage of deaths that are attributed to suicide, other external causes, and natural causes across diagnostic categories (e.g., anxiety, depression, bipolar disorder, and schizophrenia-spectrum disorder), and samples.**

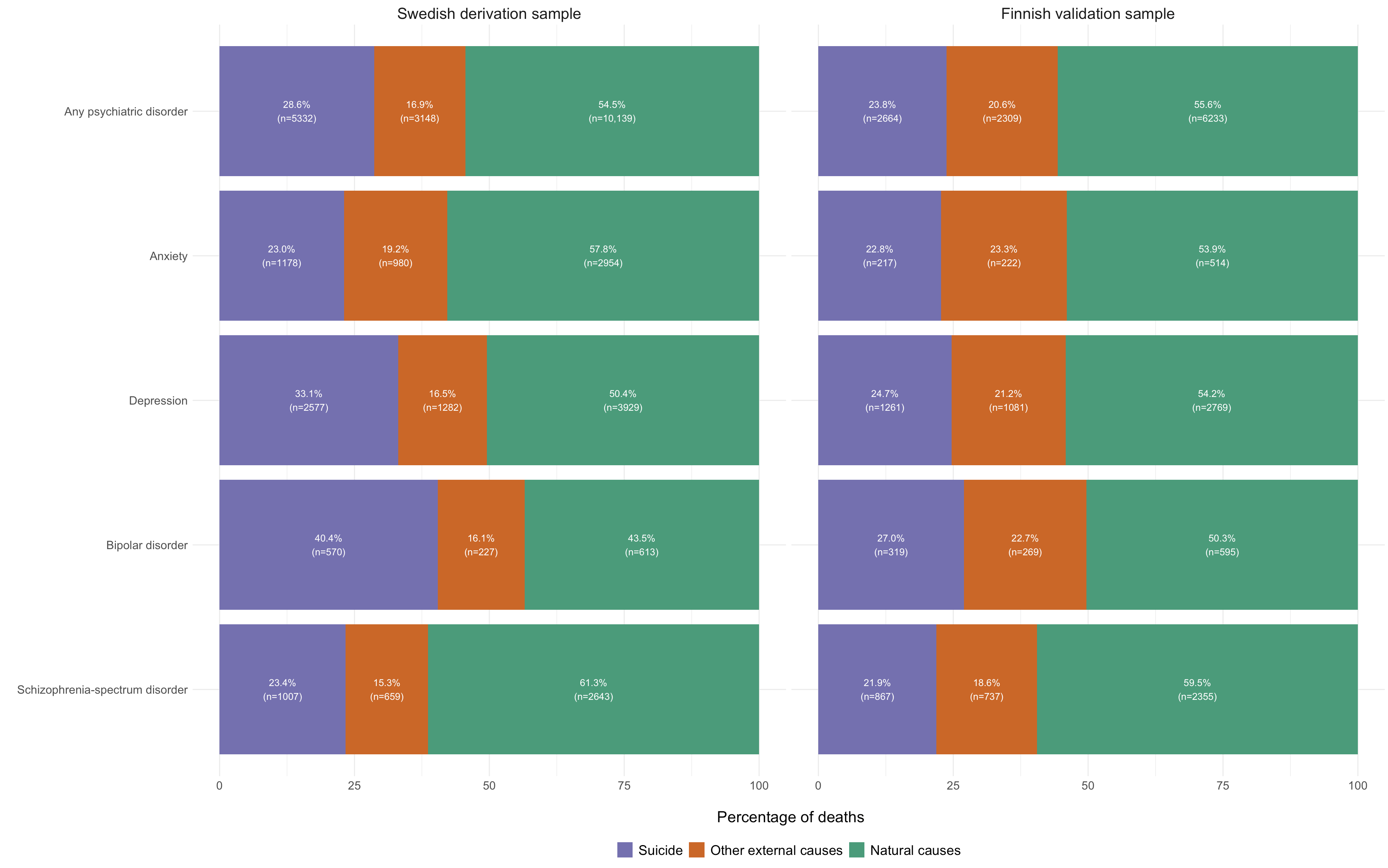

**eFigure 3. Stability plot: Individual-level predictions derived from the final model compared with 200 bootstrapped models in individuals included in the derivation sample**

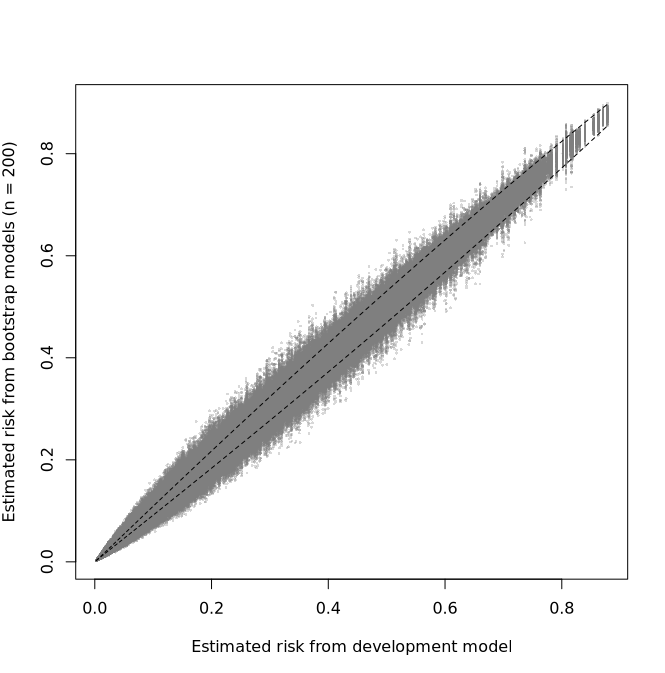

**eFigure 4. Discriminatory accuracy and calibration across Swedish counties assessed using internal-external cross-validation**

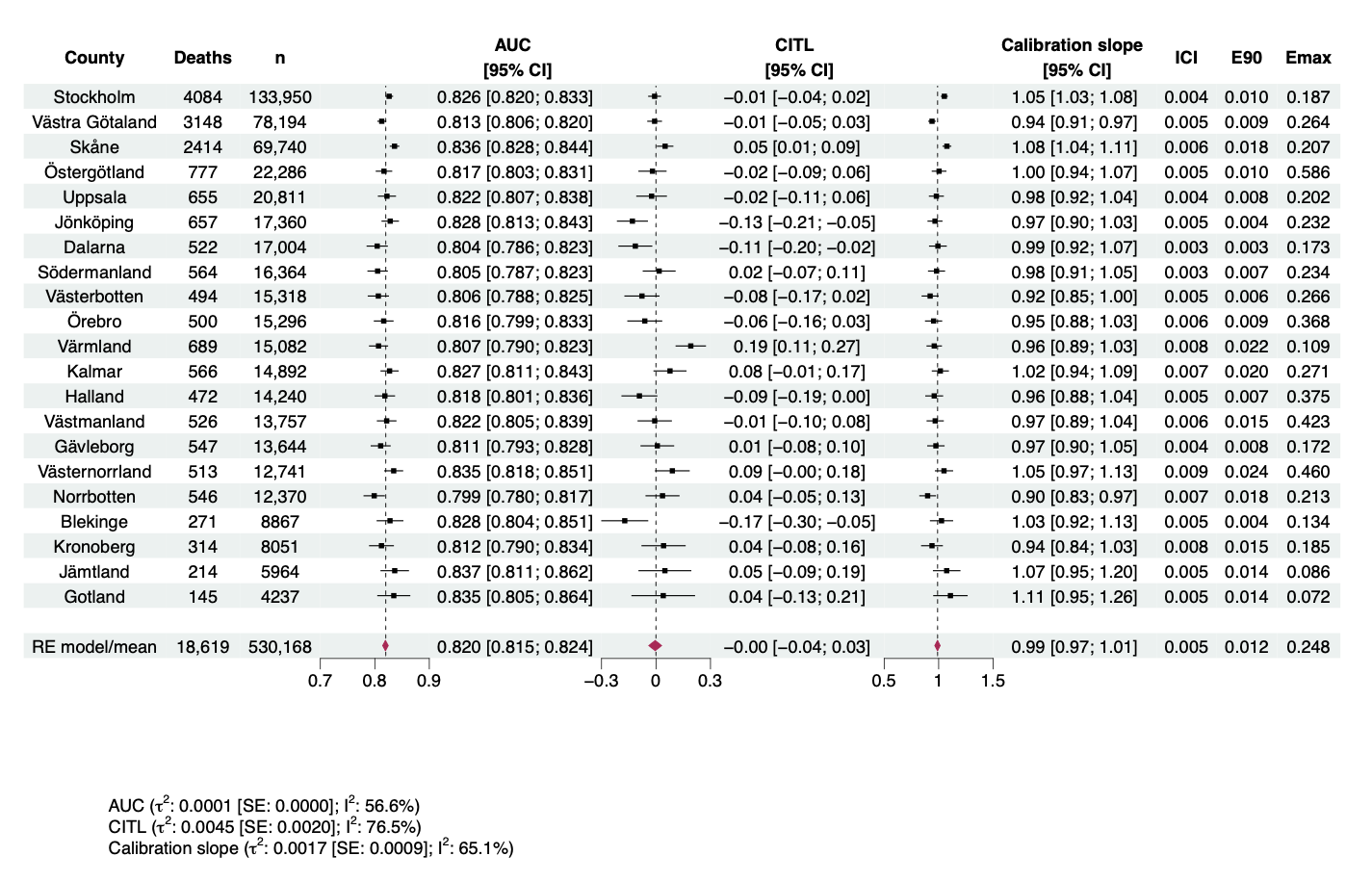

*Notes: AUC denotes the area under the receiver operator characteristic curve; CITL the calibration-in-the-large; ICI, the Integrated Calibration Index, or the weighted average of the absolute differences between the calibration curve and perfect calibration, with E90 and Emax measuring the 90th percentile and maximum value of the same differences. A total of 33 individuals lacked geographic data and were consequently excluded.*

**eFigure 5. Discriminatory accuracy and calibration in the Finnish external validation sample, including subgroup analyses geographical regions, sex, age groups, and benefit receipt**

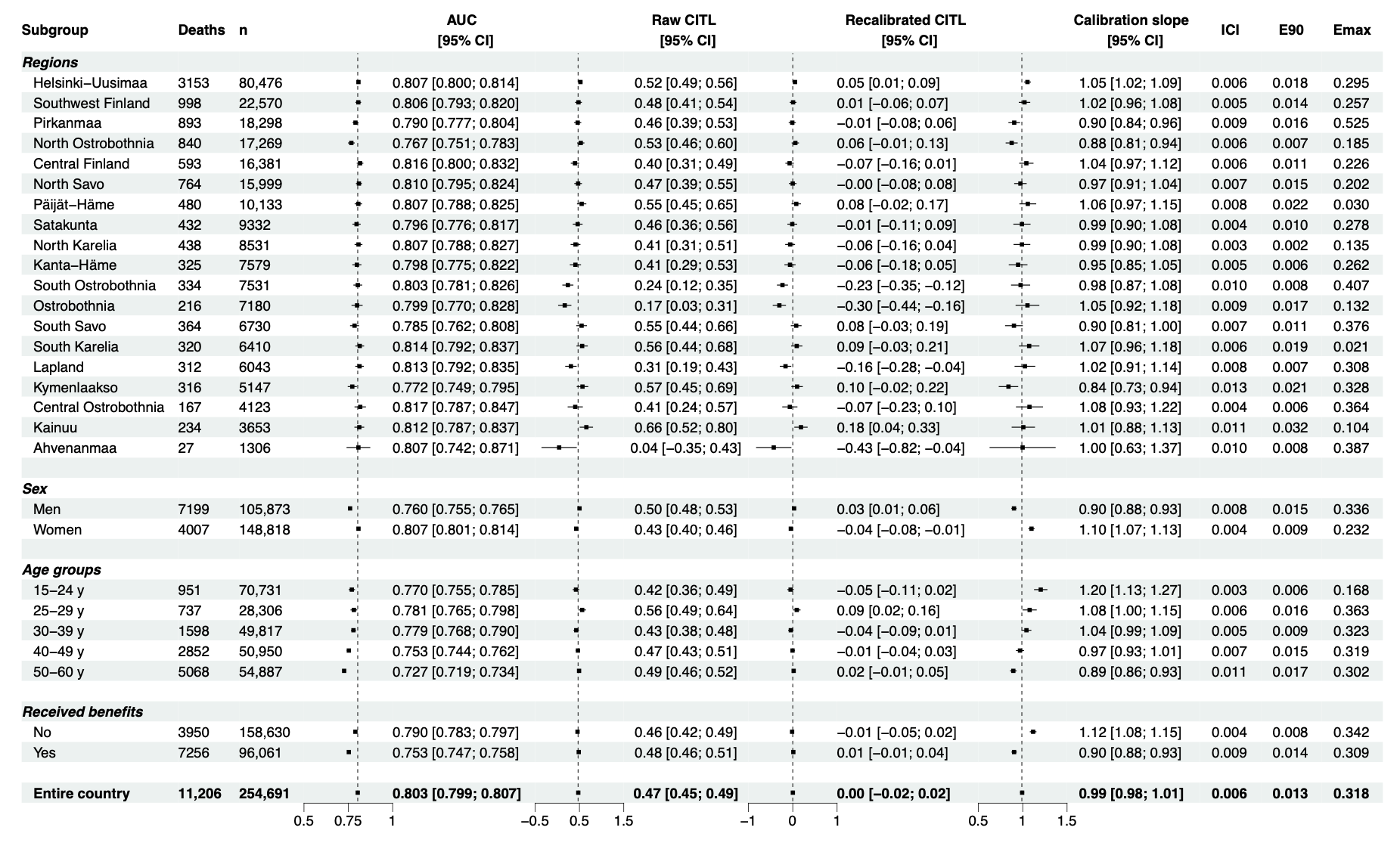

*Notes: AUC denotes the area under the receiver operator characteristic curve; CITL the calibration-in-the-large; ICI, the Integrated Calibration Index, or the weighted average of the absolute differences between the calibration curve and perfect calibration, with E90 and Emax measuring the 90th percentile and maximum value of the same differences.*

**eFigure 6. Discriminatory accuracy and calibration for post-hoc sensitivity analyses in the Finnish external validation sample**

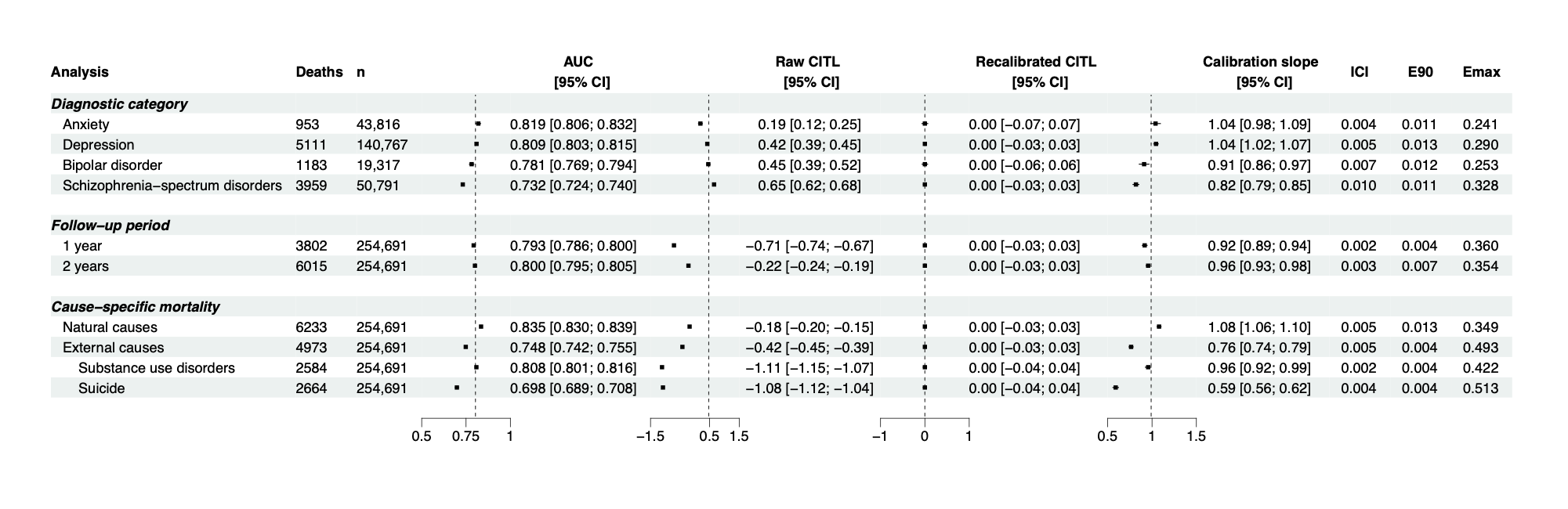

*Notes: AUC denotes the area under the receiver operator characteristic curve; CITL the calibration-in-the-large; ICI, the Integrated Calibration Index, or the weighted average of the absolute differences between the calibration curve and perfect calibration, with E90 and Emax measuring the 90th percentile and maximum value of the same differences.*
